# Clinically relevant risk threshold for predicting sudden cardiac death

**DOI:** 10.64898/2026.03.18.26348515

**Authors:** Jussi Hernesniemi, Roni Ahola, Mikko Uimonen

**Affiliations:** Faculty of Medicine and Health Technology, Tampere University, Tampere, Finland; Heart Hospital, Tampere University Hospital, Wellbeing Services County of Pirkanmaa, Tampere, Finland

**Keywords:** Sudden cardiac death, simulation, annual risk, implantable cardioverter-defibrillator

## Abstract

**Background:** There is no consensus on a risk threshold for sudden cardiac death (SCD) that could be used in practical design and evaluation of prediction models and decisions regarding implantable cardioverter-defibrillator (ICD) therapy.

**Methods:** Baseline assumptions for a simulation framework were derived from previous randomized controlled trials (n=18) to identify minimal SCD risk threshold that would translate to mortality benefit by ICD therapy also considering the effect of competing non-sudden mortality. ICD efficacy to prevent SCDs and other data for simulations were estimated using inverse-variance weighted meta-analysis of included trials. Number needed to treat (NNT) was evaluated over a five-year horizon (≤21 defined as clinically relevant).

**Results:** Correlation analysis confirmed annual SCD incidence in trial populations as the key factor associating with ICD therapy effectiveness to reduce mortality (Pearson’s r=0.653, p<0.01). In a simulation assuming 5% annual non-sudden mortality (pooled estimate of included RCTs) and a 56% (48-62%) efficacy for ICDs to reduce SCDs or similar events, 3% annual SCD risk (≈12% over five years) emerged as the lowest practical threshold even after controlling for excess (overlapping) mortality among those saved successfully from SCD by ICD therapy. The theoretical minimum threshold for annual SCD risk is 2.0%, 2.5% and 3.5% for populations with the annual incidence of non-sudden deaths 2%, 5% and 10% (assuming no overlapping mortality).

**Conclusions:** Even under substantial competing risk, a 3% annual SCD threshold appears an optimal minimum threshold for identifying patients most likely to benefit from ICD therapy if severe mortality overlap is not observed.

**Key Questions:** What is the minimal risk threshold after which ICD therapy will likely lead to meaningful reduction in overall mortality. This information is needed in practical design of clinical trials and evaluation and development of prediction models

**Key Finding:** Analysis of the data extracted from previous randomized controlled trials revealed that annual SCD risk should be at least 3% in most scenarios (with the annual incidence of non-sudden mortality ≤5%) for ICD therapy to be effective.

**Take-home Message:** Primary prevention SCD and risk models targeted to identify high-risk individual should aim for identifying patients with 3% or higher annual risk for SCD.

## INTRODUCTION

Since the publication of landmark trials decades ago showing that implantable cardioverter-defibrillation devices (ICDs) effectively lower overall mortality in patients with heart failure (HF) with reduced ejection fraction (LVEF) or in secondary prevention of sudden cardiac death (SCD)^1,2^, the medical community has struggled to found equally applicable criteria to discern reliably high-risk individuals who would benefit from ICD therapy^3^. Although SCD is still a major complication of HF^4^, even in patients with reduced LVEF the mortality benefit of ICDs seem to have declined due to decreasing rate of SCD^5,6^. The general population and patients with seemingly uncomplicated coronary artery disease in which the majority of SCDs occur remain mostly unprotected^7,8^.

Although several SCD risk models have reported favorable area under the curve values of the receiving operating characteristics curve (AUROC) values in general population^9,10^ or among patients with coronary artery disease^11^ none have been adopted in routine clinical practice. In low-event-rate populations, AUROC is heavily influenced by correct classification of low-risk individuals, whereas accurate discrimination of small high-risk subgroups is substantially more challenging^12^. This results in poor calibration in extreme-risk strata and limited generalizability especially if the model is not specifically designed to identify high risk individuals^12^.

The central paradigm in the prevention of SCD is defining the lowest practical threshold of risk at which ICD therapy becomes justified. Although this information is pivotal for risk model design and calibration and for deciding who could benefit from ICD therapy, there are no guidelines on the matter. The purpose of this meta-analysis guided simulation study is to establish clear criteria for lowest practical SCD risk threshold and to evaluate the role of other factors such as competing events due to non-sudden deaths in risk modeling.

## METHODS

This study combines meta-analytic approach to previously published randomized controlled trials (RCTs) of ICD therapy with a simulation analysis. Baseline assumptions for the simulations were obtained from the meta-analysis of previous trials. Our primary aim was to understand how SCD risk level (incidence/event rate) is associated with the success of the trial to show reduction in overall mortality by ICD therapy. The baseline incident rate was evaluated using data from the control populations of the trials (i.e. among patients who were not randomized to ICD therapy). Secondarily, we also evaluated whether other specific demographic factors (linked to SCD incidence/event rate) and the risk of competing events would associate with trial success.

Simulation study was used to search for the most optimal minimum level of annual SCD risk that could be for prediction model development and in design of RCTs to reduce mortality by ICD therapy. This risk threshold was also adjusted for the level of competing events (mortality to non-sudden deaths).

### Data search

Relevant RCTs were searched using PubMed. We included only RCTs where the allocation between ICD therapy was randomized. All trials published before 31^st^ of November 2025 were included. The results were cross-referenced to previously published literary reviews and meta-analyses based on more exhaustive literature research^1,2^. Based on the data search eighteen individual studies were included (Table 1)^13–33^. The Multicenter Unsustained Tachycardia Trial (MUSTT) was excluded due to lack of true randomization to ICD therapy^34^.

**Table 1.**
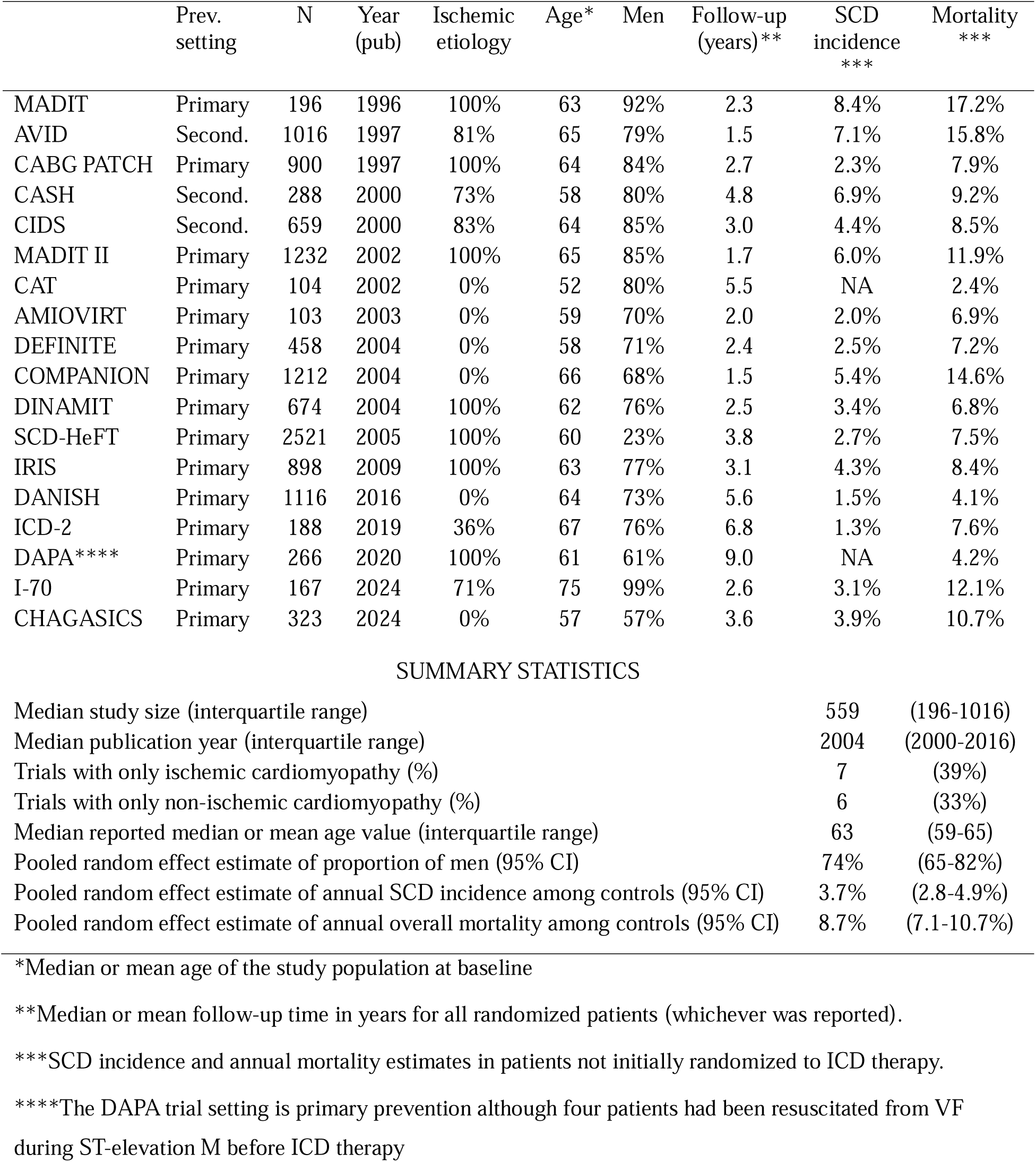
List of included randomized controlled trials and their population characteristics and approximate annual incidence of sudden cardiac deaths and annual mortality as observed in the control populations of trials. Trials have been sorted by publication year of primary endpoint analysis.

|  | Prev.<br>setting | N | Year<br>(pub) | Ischemic<br>etiology | Age* | Men | Follow-up<br>(years)** | SCD<br>incidence<br>*** | Mortality<br>*** |
| --- | --- | --- | --- | --- | --- | --- | --- | --- | --- |
| MADIT | Primary | 196 | 1996 | 100% | 63 | 92% | 2.3 | 8.4% | 17.2% |
| AVID | Second. | 1016 | 1997 | 81% | 65 | 79% | 1.5 | 7.1% | 15.8% |
| CABG PATCH | Primary | 900 | 1997 | 100% | 64 | 84% | 2.7 | 2.3% | 7.9% |
| CASH | Second. | 288 | 2000 | 73% | 58 | 80% | 4.8 | 6.9% | 9.2% |
| CIDS | Second. | 659 | 2000 | 83% | 64 | 85% | 3.0 | 4.4% | 8.5% |
| MADIT II | Primary | 1232 | 2002 | 100% | 65 | 85% | 1.7 | 6.0% | 11.9% |
| CAT | Primary | 104 | 2002 | 0% | 52 | 80% | 5.5 | NA | 2.4% |
| AMIOVIRT | Primary | 103 | 2003 | 0% | 59 | 70% | 2.0 | 2.0% | 6.9% |
| DEFINITE | Primary | 458 | 2004 | 0% | 58 | 71% | 2.4 | 2.5% | 7.2% |
| COMPANION | Primary | 1212 | 2004 | 0% | 66 | 68% | 1.5 | 5.4% | 14.6% |
| DINAMIT | Primary | 674 | 2004 | 100% | 62 | 76% | 2.5 | 3.4% | 6.8% |
| SCD-HeFT | Primary | 2521 | 2005 | 100% | 60 | 23% | 3.8 | 2.7% | 7.5% |
| IRIS | Primary | 898 | 2009 | 100% | 63 | 77% | 3.1 | 4.3% | 8.4% |
| DANISH | Primary | 1116 | 2016 | 0% | 64 | 73% | 5.6 | 1.5% | 4.1% |
| ICD-2 | Primary | 188 | 2019 | 36% | 67 | 76% | 6.8 | 1.3% | 7.6% |
| DAPA**** | Primary | 266 | 2020 | 100% | 61 | 61% | 9.0 | NA | 4.2% |
| I-70 | Primary | 167 | 2024 | 71% | 75 | 99% | 2.6 | 3.1% | 12.1% |
| CHAGASICS | Primary | 323 | 2024 | 0% | 57 | 57% | 3.6 | 3.9% | 10.7% |

SUMMARY STATISTICS
|  |  |  |
| --- | --- | --- |
| Median study size (interquartile range) | 559 | (196-1016) |
| Median publication year (interquartile range) | 2004 | (2000-2016) |
| Trials with only ischemic cardiomyopathy (%) | 7 | (39%) |
| Trials with only non-ischemic cardiomyopathy (%) | 6 | (33%) |
| Median reported median or mean age value (interquartile range) | 63 | (59-65) |
| Pooled random effect estimate of proportion of men (95% CI) | 74% | (65-82%) |
| Pooled random effect estimate of annual SCD incidence among controls (95% CI) | 3.7% | (2.8-4.9%) |
| Pooled random effect estimate of annual overall mortality among controls (95% CI) | 8.7% | (7.1-10.7%) |

\*Median or mean age of the study population at baseline
\*\*Median or mean follow-up time in years for all randomized patients (whichever was reported).
\*\*\*SCD incidence and annual mortality estimates in patients not initially randomized to ICD therapy.
\*\*\*\*The DAPA trial setting is primary prevention although four patients had been resuscitated from VF during ST-elevation M before ICD therapy

### Data Extraction

Information of the general setting of the trial, trial population (general features) and ICD therapy success on reducing overall mortality and sudden cardiac deaths (or similarly defined endpoints such as sudden arrhythmic death) was extracted. Individual participant data (IPD) was not used. Main baseline population characteristics such as median age (or mean age if median was not reported), sex distribution, study setting (secondary or primary prevention od SCD) was recorded. Incidence of follow-up events (sudden cardiac deaths or similar events such as sudden arrhythmic events, non-sudden deaths or non-arrhythmic deaths and overall mortality) and duration of follow-up was recorded separately by treatments arms (stratification by intention to treat). No attempt was made to control for possible cross-over between treatment arms due to lack of specific information in each trial in relation to the outcomes. The data from main reports were used for each study in the analysis (i.e. including only the main publication addressing the primary hypothesis for the first time) unless specified otherwise.

### Statistical analysis

Annual incidence of SCDs was estimated from study-level event counts and follow-up duration in the control populations of the trials. Person-time was approximated as cohort size multiplied by reported median follow-up, and incidence rates were calculated as events per person-year. Incidence rates were converted to annual risk, assuming constant hazards using an exponential model.

The effects of ICD therapy on all-cause mortality and SCD were pooled using inverse-variance weighted meta-analysis of log-transformed hazard ratios (HRs) assuming fixed (Significant heterogeneity was not observed by assessing the Q statistic, I², and τ² and tested p-value for heterogeneity). Incidence rates for SCD, competing non-sudden mortality and overall mortality across all trials were also evaluated by a pooled inverse-variance meta-analysis. The association between different study characteristics (annual incidence of SCDs, annual mortality proportion of SCDs of off deaths, and baseline population median/mean age, sex distribution, proportion of patients with ischemic cardiomyopathy and publication year) was assessed using correlation analysis of point estimates and additionally by controlling for uncertainty in the treatment-effect estimates (observed mortality reduction) by inverse-variance weighting.

A model-based simulation framework was used to evaluate the impact of baseline SCD incidence on ICD effectiveness for a fixed five-year period after ICD implantation. This time window was selected for practical reasons: It exceeds the follow-up times of most ICD trials (see results) but stays below the expected battery life of single ICD device and because the same time window is recommended to be used to evaluate ICD need in patients with HCM and LQT syndrome^35,36^.

Under an exponential survival model, annual incidences of SCD and non-sudden death were converted to cause-specific hazards. All-cause mortality hazard was modeled as the sum of the SCD hazard and the non-sudden death hazard. ICD therapy was assumed to act only on the SCD hazard, modeled as a proportional reduction according to the meta-analyzed hazard ratio. Hazard ratio and number needed to treat (NNT) were calculated. Uncertainty was incorporated using Monte Carlo simulation (20,000 iterations). Between-study heterogeneity was modeled on the log-hazard-ratio scale using a normal distribution with standard deviation equal to the estimated heterogeneity parameter (τ). In addition, estimation uncertainty in the pooled treatment effect was incorporated by sampling the mean log-hazard ratio from a normal distribution with variance derived from the 95% confidence interval of the meta-analyzed effect estimate. Dependence between sudden cardiac death (SCD) and non-sudden mortality was explored in a sensitivity analysis by allowing a proportion of ICD-prevented SCD events to be followed by a replacement non-sudden death within a prespecified 0- to 12-month window, assuming a uniform distribution of event timing within that interval. The proportion of displaced events was simulated using a replacement parameter ranging from 0 to 1, and the number needed to treat (NNT) was recalculated. Based on the results of seminal trials showing ICD treatment to reduce mortality, an NNT of ≤21as observed in seminal trials showing ICD therapy to reduce mortality was considered as clinically relevant threshold for ICD treatment to be effective^13–16,23^. All analyses were performed in R software, with two-sided P values <0.05 considered statistically significant.

## RESULTS

### Correlation between trial characteristics and ICD effectiveness to reduce overall mortality

Eighteen RCTs with 12,321 patients were included in the meta-analysis. The median follow-up time in the eighteen RCTs was 2.8 years (interquartile range between 2.2 – 4.9 years) during which 935 patients suffered an SCD, or a similar event and overall 2,933 patients died (any cause). Details of these studies are presented in Table 1. Direct comparison of the point estimates of ICD effectiveness for reducing overall mortality and different trial characteristics showed that annual incidence of SCDs and annual overall mortality were the only statistically significant factors associated with ICD therapy effectiveness (Pearson’s r -0.653, p=0.006 for SCD incidence and Pearson’s r -0.585, p=0.017 for overall mortality incidence, Table 2). Notably, of the top five trials with highest annual incidence of SCD among controls ranging between 8.4-5.4% (Table 1), three reported statistically significant (p<0.05) reduction in overall mortality (MADIT, AVID and MADIT II) and two other non-significant but consistent results (i.e. similar direction of the effect in CASH and Companion trials) (Figure 1). The precision weighted correlation analysis showed a similar direction and magnitude of association (r = −0.59, p = 0.017)(Figure 2). Proportion on SCDs of all deaths, sex, distribution, proportion of patients with ischemic cardiomyopathy or publication year were not statistically significantly associated with ICD therapy effectiveness in simple correlation analysis although some of the correlation coefficients were notable (p=NS for all, Table 2)

**Figure 1.**
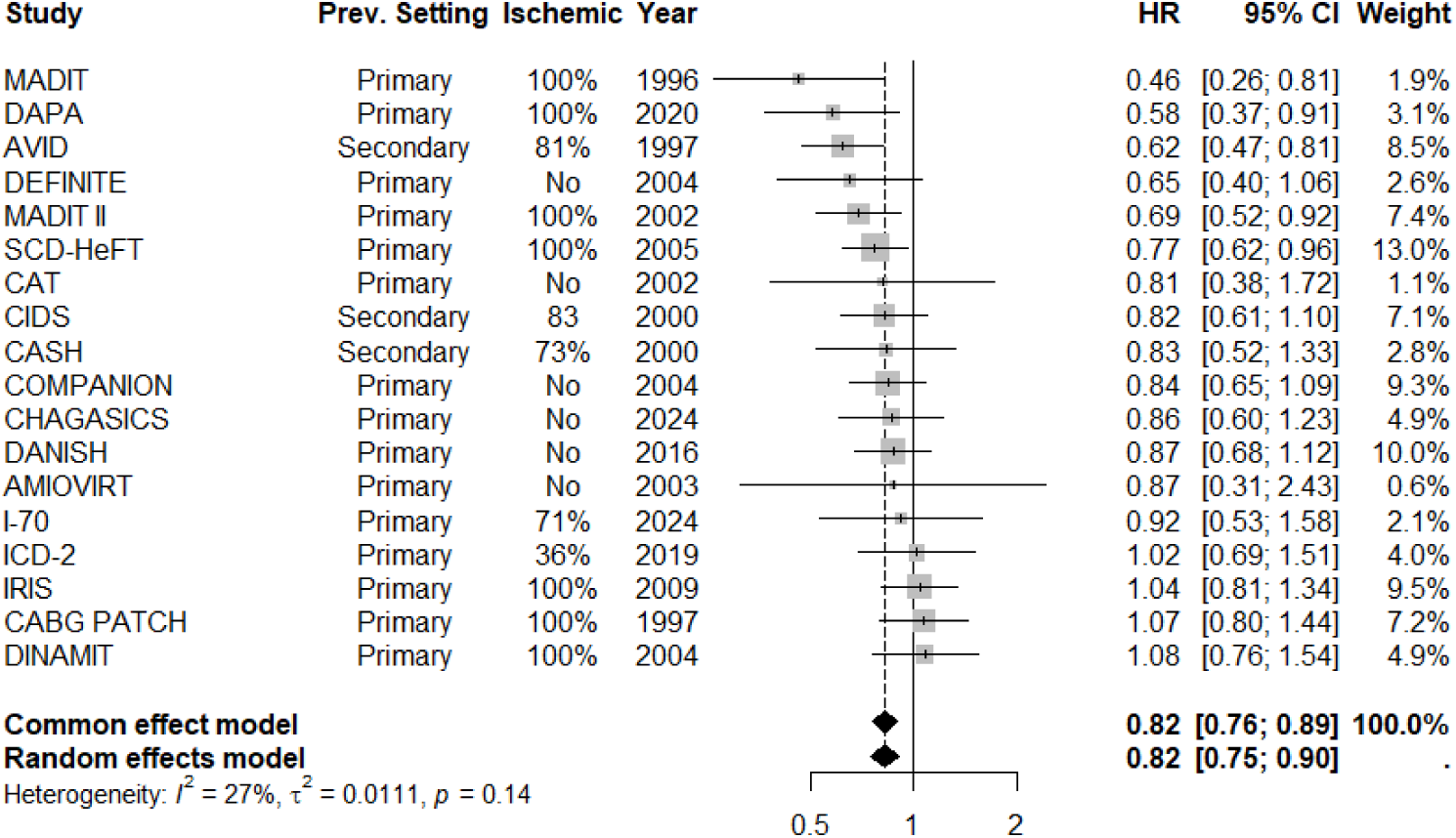
Meta-analysis of the effect of ICD therapy on overall mortality in randomized controlled clinical trials. Studies are sorted according to the observed efficacy of ICD.

**Figure 2.**
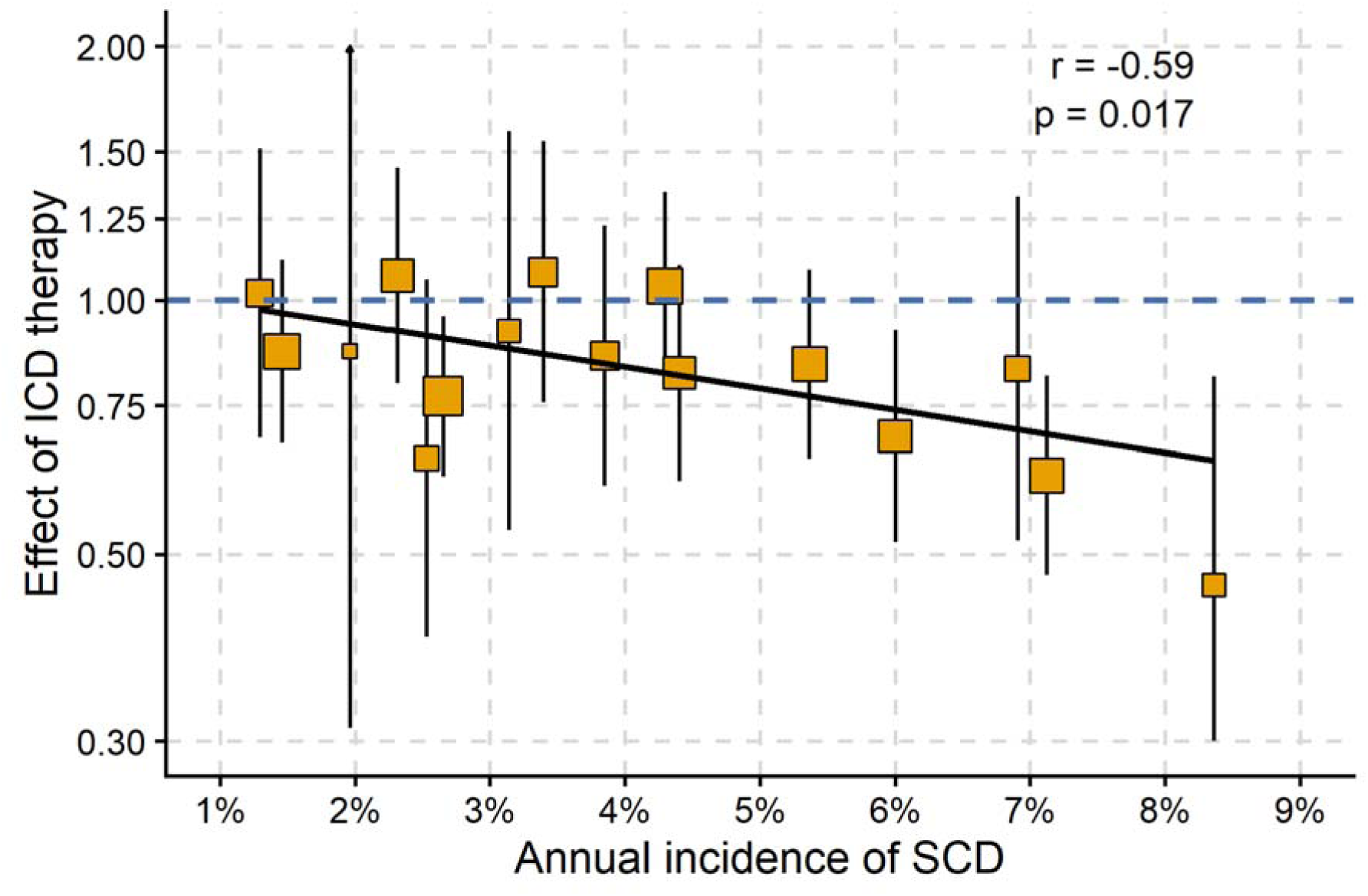
Precision weighted correlation analysis between annual incidence of sudden cardiac death or similar events observed in control populations of randomized controlled trials (x-axis) and the observed effect of ICD therapy for reducing mortality (y -axis). The effect of ICD therapy is presented in study specific hazard ratios and 95% confidence intervals.

**Table 2.** Correlation coefficients between different population characteristics and observed mortality benefit for ICD therapy in randomized controlled trials. The correlation was calculated between the point estimates of listed characteristics and beta-value for mortality reduction calculated from the published hazard ratio values.

|  | Pearsons r | P-value |
| --- | --- | --- |
| Annual incidence of sudden cardiac death* | -0.650 | 0.006 |
| Annual mortality (any cause) | -0.629 | 0.009 |
| Proportion of sudden cardiac deaths of all deaths* | -0.249 | 0.352 |
| Proportion of men | -0.065 | 0.810 |
| Proportion of patients with ischemic cardiomyopathy | 0.311 | 0.241 |
| Publication year | 0.422 | 0.104 |
| Median/mean age of study population | 0.137 | 0.614 |

\*The Defibrillator After Primary Angioplasty (DAPA) trial and the Cardiomyopathy Trial (CAT) not included in the analysis due to missing or uncertain data.

### Simulation of the ICD therapy effectiveness

In meta-analysis, ICDs reduced SCDs or similar events by 56% (HR 0.44, 0.38-0.52, p < 1 × 10C^24^ for fixed effect model, I^2^=7%, p=0.37 for heterogeneity) (Figure 3) in population with pooled estimate of 5.1% (95% CI 4.8-5.4%) for annual incidence of non-sudden death. Using these as baseline assumptions, simulated the relative risk reduction and NNT to save one life across varying annual SCD incidence rates. The gradual decrease in relative risk of death by ICD therapy at different levels of SCD incidence is shown in Figure 4. In terms of NNT, the function seems non-linear showing that from 0 baseline to <2% annual incidence of SCD, the NNT-values improve rapidly (decrease) and after 2.0% annual incidence of SCD (NNT of 25 with 95% CI 22-28), the improvement slows down reaching an NNT of 9 (8-11) at 6% annual incidence and finally plateauing at NNT values closer to 5 at higher annual incidence values (Figure 5). The HR and NNT values for different values of annual incidence of SCD and at this 5% fixed level of mortality due to non-sudden causes (competing events) are presented in Table 3. Based on the simulation, in a population with 5% fixed non-sudden death incidence, an annual incidence of 2.5% for SCD would be sufficient to show clinically relevant benefit for ICD therapy (NNT of 20 with 95% CI 18-23, corresponding HR 0.81, 95 CI 0.79-0.84). For reference, in meta-analysis of previous RCTs, the pooled estimate of SCD incidence was 3.7% (2.8-4.9%) and the HR value associated with ICD therapy effectiveness to reduce overall mortality was 0.82 (0.77-0.90, p < 1 × 10C^5^)(Figure 1.).

**Figure 3.**
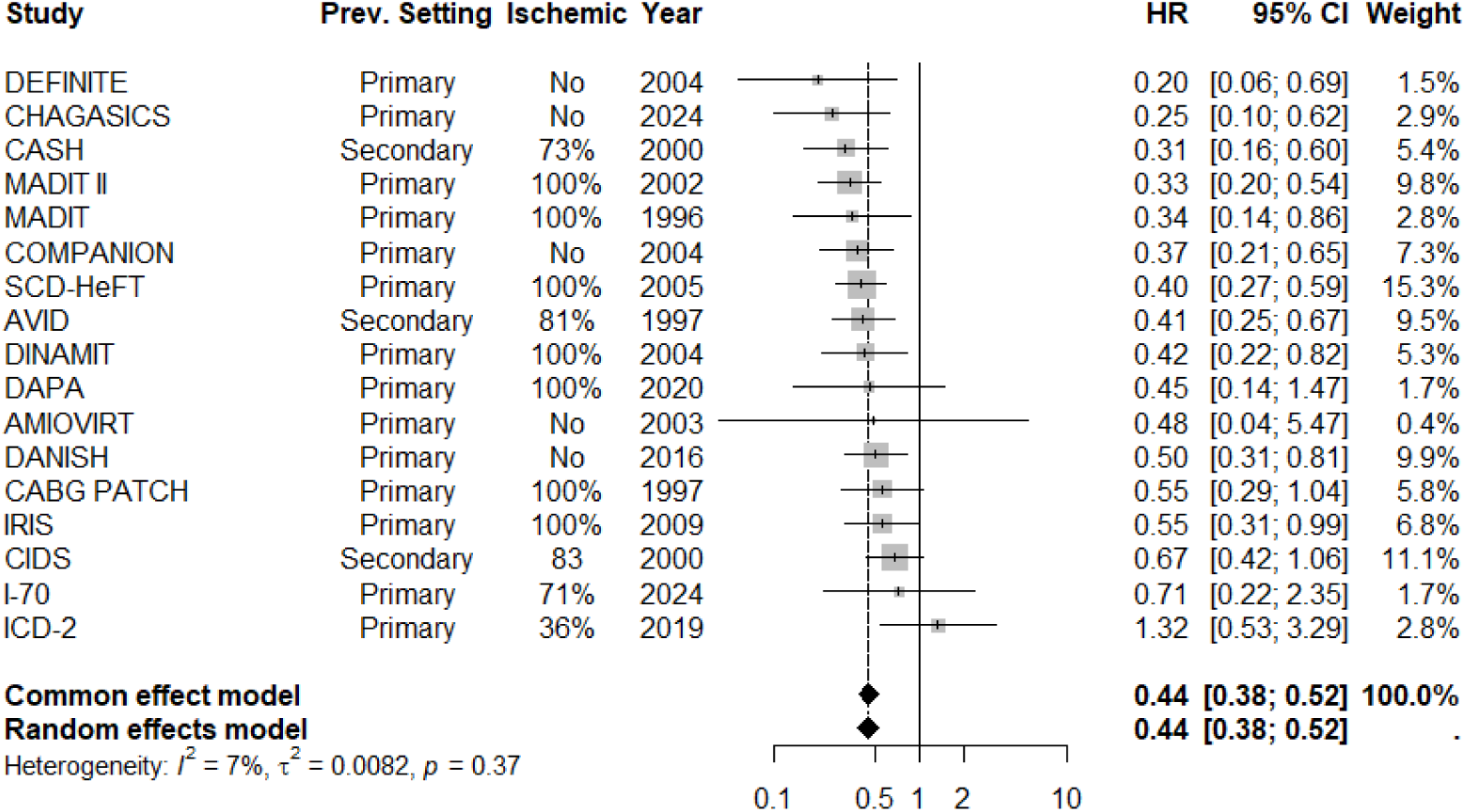
Meta-analysis of the effect of ICD therapy to reduce sudden cardiac deaths or similar events in randomized controlled clinical trials. Studies are sorted according to the observed effect of ICD therapy.

**Figure 4.**
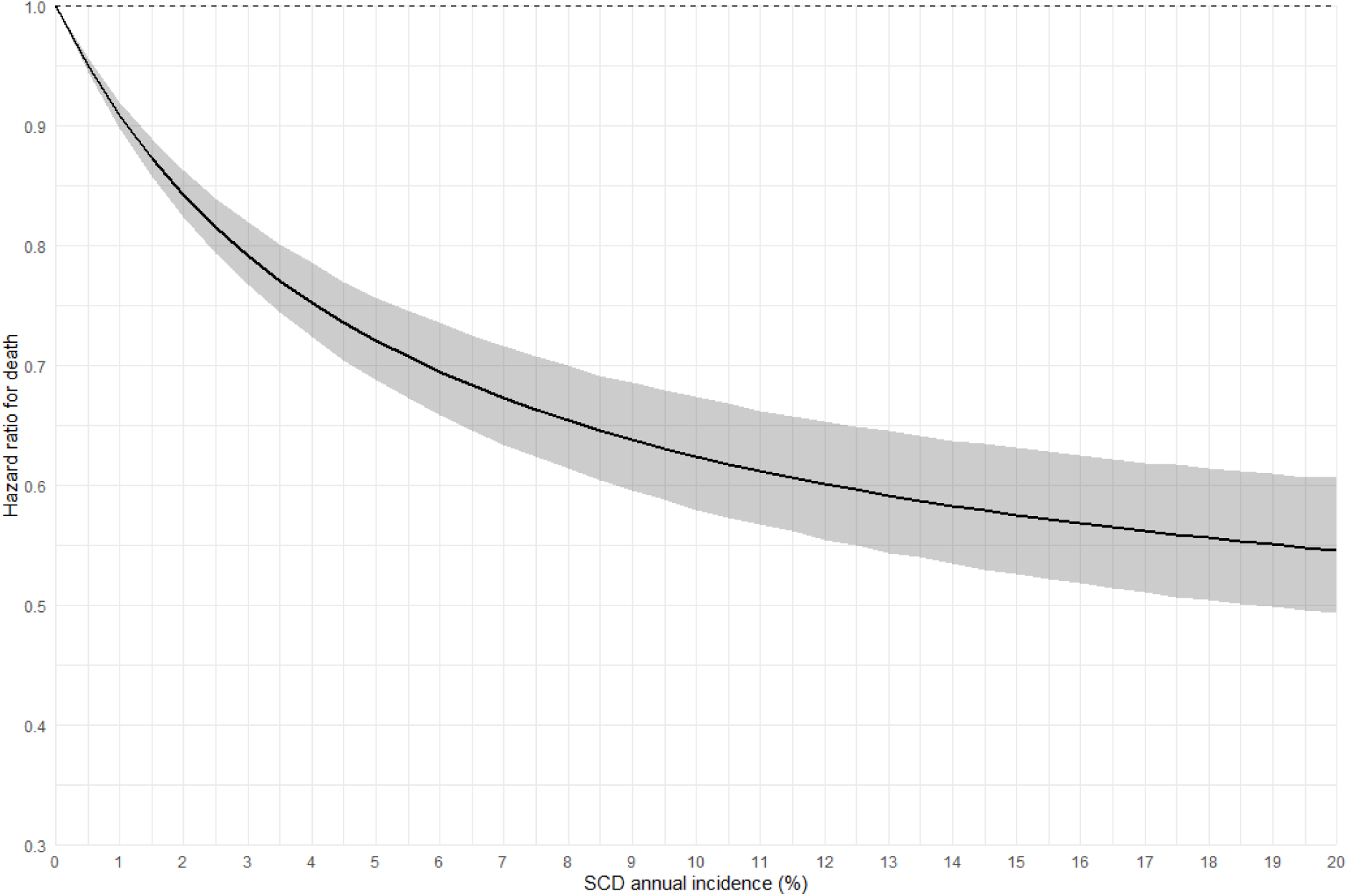
Simulation of the relative mortality benefit of ICD therapy across varying annual SCD incidence rates, assuming a constant annual non-SCD mortality of 5%.

**Figure 5.**
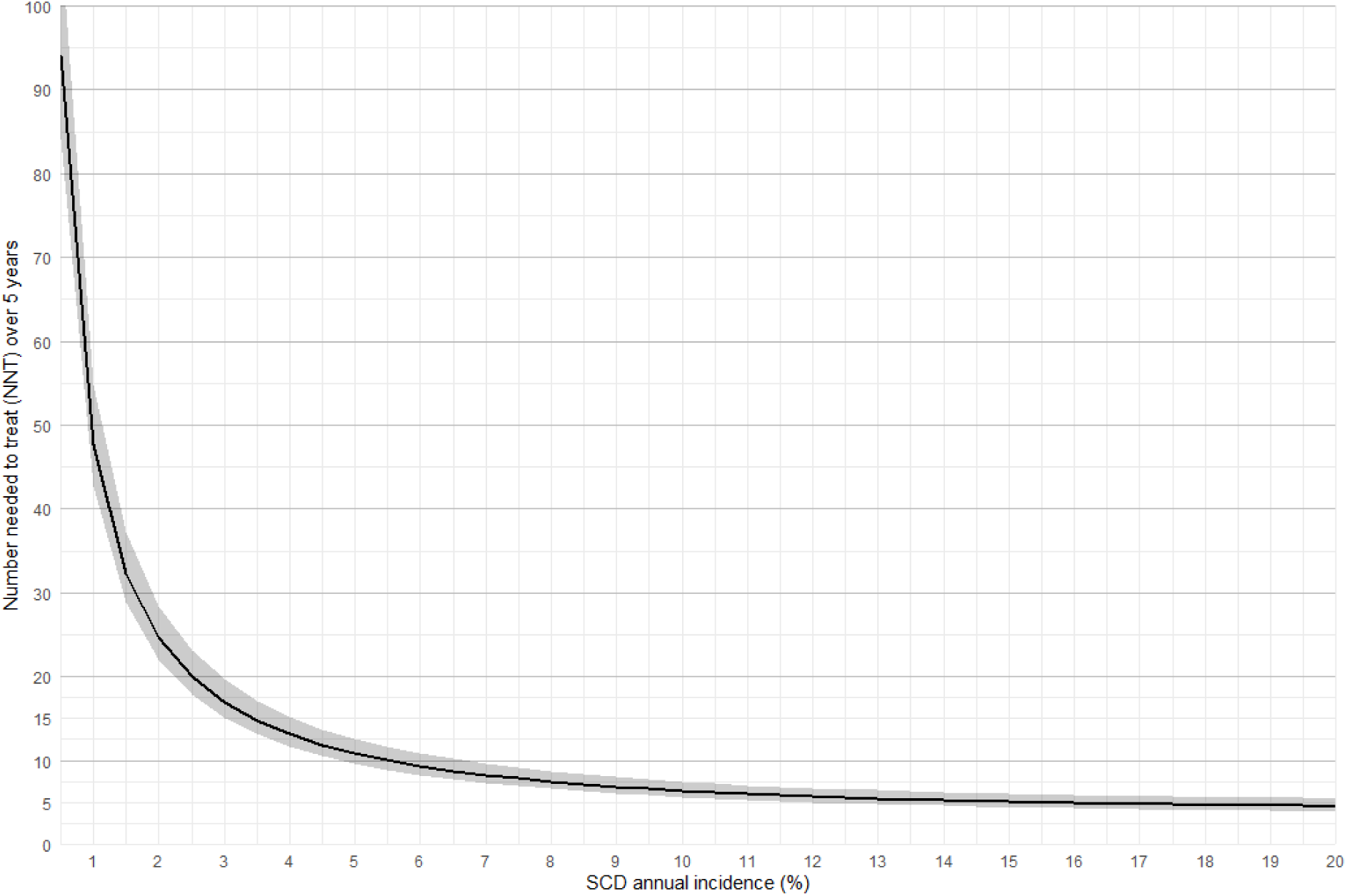
Simulation of the 5-year number needed to treat (NNT) to prevent one death with ICD therapy across varying annual SCD incidence rates, assuming constant annual non-SCD mortality of 5%.

**Table 3.** Simulation of the relation between annual incidence of sudden cardiac death and overall mortality reduction achievable by ICD therapy in different scenarios with 5% fixed estimate for annual incidence of non-sudden deaths.

| SCD incidence | Hazard ratio | 95% CI | Number needed to treat | 95% CI |
| --- | --- | --- | --- | --- |
| 1.0% | 0.91 | 0.90-0.91 | 48 | 42-55 |
| 1.5% | 0.87 | 0.86-0.89 | 32 | 29-37 |
| 2.0% | 0.84 | 0.82-0.86 | 25 | 22-28 |
| 2.5% | 0.82 | 0.79-0.84 | 20 | 18-23 |
| 3.0% | 0.79 | 0.77-0.82 | 17 | 15-20 |
| 3.5% | 0.77 | 0.74-0.80 | 15 | 13-17 |
| 4.0% | 0.75 | 0.72-0.79 | 13 | 12-15 |
| 4.5% | 0.74 | 0.70-0.77 | 12 | 11-14 |
| 5.0% | 0.72 | 0.69-0.76 | 11 | 10-12 |
| 5.5% | 0.71 | 0.67-0.75 | 10 | 9-12 |
| 6.0% | 0.69 | 0.66-0.74 | 9 | 8-11 |

### Sensitivity analyses

For sensitivity analyses, we also analyzed what would be an appropriate limit for annual incidence of SCD in populations with different mortality to non-sudden causes. Assuming only a 2% annual risk of competing events, a clinically relevant limit for annual incidence of SCD would be 2% (NNR of 21 with 95% CI 19-24) and assuming an extreme 10% annual risk of competing events, a clinically relevant limit for annual incidence of SCD would be 3.5% (NNT of 19 with 95% CI 17-22). Additional analysis was performed accounting for possible misevaluation of non-sudden death risk in patients who ultimately suffer an SCD event (i.e. SCD victims might have manifestly higher risk of dying regardless whether their SCD was saved or not by ICD therapy). If the significant SCD risk threshold is assumed to be 2.5% and the baseline non-sudden death risk is fixed to 5%, only ≤4.3% of SCD could be followed by a replacement event within a one year to achieve NNT≤21. Setting significant SCD risk threshold to 3% would allow the replacement probability to be 17.3% and with a threshold of 3.5%, the replacement probability could be up to 26% to achieve NNT≤21.

## DISCUSSION

The results of this meta-analysis and simulation study highlight the annual incidence of SCD as a key metric in the development of risk prediction models and in evaluating the potential benefit of ICD therapy for primary prevention. When the annual incidence of SCD is ≥2.5%, ICD therapy leads to clinically relevant reduction in overall mortality in populations with an annual competing mortality ≤5% if no overlap is observed between SCD and non-sudden death risk. However, a 3% threshold allows for some overlap in risk estimation and is likely a more practical choice.

The incidence of SCD in any population is inherently determined by disease-specific characteristics and other predisposing factors such as age or other comorbidities. An ideal risk factor would be very specific only to SCD. Age is also associated with the risk of competing events (non-sudden deaths) which can alter, to some extent, the possible benefit of ICD therapy as seen in individual participant data analysis of RCTs^37^. Our trial-level meta-regression did not detect that factor such as population age, sex distribution or aetiology of HF (ischemic or non-ischemic) across the trials would significantly modify the effectiveness of ICD therapy, observations which are in line with previous meta-analysis^38^. However, aggregate-data meta-regression has limited statistical power due to low between-study variability in covariate distributions and small numbers of included studies.

In populations with low ratio between SCD and non-sudden deaths, the benefit of ICD therapy may be attenuated. This likely explains the neutral results of the I-70 trial (primary prevention in the elderly)^25^ and ICD2 (primary prevention in dialysis patients)^24^, in which ICD therapy did not reduce all-cause mortality. This limitation is truly exacerbated if the incidence of SCD is not independent of incidence of non-sudden deaths. Preventing SCD will not reduce overall mortality if patients are more likely to die from non-sudden causes within a relatively short time frame after an adequate (and successful) ICD therapy for life-threatening ventricular arrhythmia. This mechanism was perfectly illustrated in the IRIS, DINAMIT and CABG PATCH trials, where ICD therapy failed to improve survival despite reducing successfully sudden arrhythmic events due to complete replacement of saved sudden deaths for non-arrhythmic deaths during the follow-up^26,27,39^.

For simplicity, we assumed independence between the cause-specific hazards for SCD and non-sudden death, which is unlikely to hold entirely in real-world populations with older or multimorbid patients. Unfortunately, the true extent of replacement events can be generally only tested after ICD therapy is implemented. According to our simulation, if 3% annual SCD incidence is assumed as a clinically relevant threshold and 5% incidence of non-sudden deaths is assumed (corresponding to the pooled incidence of previous RCTs), up to 17% of patients saved by ICD therapy could still die within a year of the event non-suddenly with NNT values maintained at 21 or less, which was set as a benchmark based on the results of seminal trials of ICD efficacy^13–16,23^. Due to the difficulty predicting this overlap, we recommend a conservative strategy prioritizing SCD-specific risk factors and using sub-distribution hazard models, which account for the association between putative risk factors and competing events^40^. Unfortunately, most traditional clinical risk factors are strongly associated with the overlapping risk of both SCD and non-sudden death, which explains their limited discrimination ability^3^. Alternatively, risk modelling using fixed and sufficiently long-time horizons (for example 5-year risk) may improve clinical applicability.

Accepting the confounding by overlapping risk, an annual SCD incidence of approximately ∼ 3% represents the lowest practical threshold at which primary-prevention ICD therapy is likely to provide clinically meaningful benefit. This limit is also close to the pooled estimate of SCD incidence in all included ICD trials (3.7%). Over a fixed 5-year horizon, the three percent annual SCD incidence corresponds to a cumulative SCD risk of 12.4% in an optimistic scenario in which causes of death are independent (non-sudden death annual incidence assumed to be 5%). In hypertrophic cardiomyopathy (HCM), current European guidelines recommend ICD implantation should be considered at a ≥6% 5-year SCD risk (HCM Risk-SCD model)^36^, while in long QT syndrome (LQTS) a ≥5% 5-year risk threshold has been proposed (1-2-3-LQT risk calculator) for asymptomatic patients. These thresholds are based on an optimistic assumption of complete prevention of SCD with ICD therapy, yielding theoretical NNTs of approximately 13 for HCM and 9 for LQTS^35,36^. Assuming a relative mortality reduction of ∼56% with ICD therapy instead of 100% efficacy (as in these publications), the NNT values would be approximately doubled aligning them closer with our simulation-based estimates.

The estimated NNT is also influenced by the selected time horizon. We used a fixed 5-year window, although follow-up durations in several randomized ICD trials have been shorter. Under a constant risk and constant effect assumption, a shorter time horizon would yield a higher NNT. In practice, however, SCD risk may be concentrated early in follow-up in some patient groups, meaning that a shorter time horizon could still capture a large share of treatment benefit. For this reason, a shorter time horizon would probably be useful as well, if the proportion of SCDs reaches high enough level (for example the 12% limit corresponding to 3% annual incidence over a five-year period).

In conclusion, we propose 3% annual incidence of SCD (or overall ∼12%) over five-years as a pragmatic benchmark for clinically meaningful SCD risk in classifiers designed to identify individuals who may benefit from preventive ICD therapy.

## ACKNOWLEDGEMENTS

None.

## ETHICS STATEMENTS

This study was based on a meta-analysis of previously published summary data from randomized controlled trials. No individual participant level data was used.

## DATA AVAILABILITY STATEMENT

All analyses of this work are based on previously published summary statistics of randomized controlled trials.

